# Machine learning predicts metastatic progression using novel differentially expressed lncRNAs as potential markers in pancreatic cancer

**DOI:** 10.1101/2023.11.01.23297724

**Authors:** Hasan Alsharoh

## Abstract

Pancreatic cancer (PC) is associated with high mortality overall. Recent literature has focused on investigating long noncoding RNAs (lncRNAs) in several cancers, but studies on their functions in PC are lacking. The purpose of this study was to identify novel lncRNAs and utilize machine learning to techniques to predict metastatic cases of PC using the identified lncRNAs. To identify significantly altered expression of lncRNA in PC, data was collected from The Cancer Genome Atlas (TCGA) and RNA-sequencing (RNA-seq) transcriptomic profiles of pancreatic carcinomas were extracted for differential gene expression analysis. To assess the contribution of these lncRNAs to metastatic progression, different ML algorithms were used, including logistic regression (LR), support vector machine (SVM), random forest classifier (RFC) and eXtreme Gradient Boosting Classifier (XGBC). To improve the predictive accuracy of these models, hyperparameter tuning was performed, in addition to reducing bias through the synthetic minority oversampling technique. Out of 60,660 gene transcripts shared between 151 PC patients, 38 lncRNAs that were significantly differentially expressed were identified. To further investigate the functions of the novel lncRNAs, gene set enrichment analysis (GSEA) was performed on the population lncRNA panel. GSEA results revealed enrichment of several terms implicated in proliferation. Moreover, using the 4 ML algorithms to predict metastatic progression returned 76% accuracy for both SVM and RFC, explicitly based on the novel lncRNA panel. To the best of my knowledge, this is the first study of its kind to identify this lncRNA panel to differentiate between non-metastatic PC and metastatic PC, with many novel lncRNAs previously unmapped to PC. The ML accuracy score reveals important involvement of the detected RNAs. Based on these findings, I suggest further investigations of this lncRNA panel *in vitro* and *in vivo*, as they could be targeted for improved outcomes in PC patients, as well as assist in the diagnosis of metastatic progression based on RNA-seq data of primary pancreatic tumors.

## 1. Introduction

Pancreatic cancer (PC) is one of the deadliest cancers, with an overall five-year survival between 7.2 and 10% according to the literature ^1,2^. Evidence suggests that PC is often diagnosed until the late stages of tumorigenesis, likely contributing to its high mortality rate ^3^. Recent literature has provided increasing evidence regarding the involvement of long noncoding RNAs (lncRNAs) in the development, invasiveness, angiogenic potential, chemotherapeutic resistance and metastatic capacity of PC ^4^.

LncRNAs are RNA molecules characterized by having an arbitrary lower cutoff of 200 nucleotides that have been shown not to code for proteins post-transcriptionally ^4,5^. LncRNAs have been shown to play complex roles in biological processes in various tissues, with possible implications in DNA repair, cellular proliferation, and human diseases, which made them a common target for recent literature to investigate in cancer ^6^. lncRNAs have further been used as biomarkers for overcoming chemoresistance, as well as for the diagnosis of several cancers, including PC ^7–10^.

Emerging research has been able to provide evidence regarding the use of lncRNAs for improved diagnostic accuracy, prognosis prediction, and treatment adjustment using various methods, including machine learning (ML) techniques ^8–10^. Literature regarding the utilization of ML algorithms has been rapidly rising, with literature urging more rapid use of such algorithms in oncology to increase diagnostic accuracy or to further improve on the available algorithms ^11–13^.

The aim of this study was to investigate potential lncRNAs involved in the metastatic progression of PC based on RNA-sequencing (RNA-seq) data. To achieve this objective, publicly available data from the cancer genome atlas (TCGA) for 172 patients was collected, and the data was filtered according to predefined inclusion and exclusion criteria, which resulted in 151 PC records. PC records were further categorized according to their TNM staging, and tumor data were separated into tumors with metastatic activity (TMAs) and tumors without metastatic activity (TWAs). Using bioinformatics analytic techniques, I identified 125 differentially expressed transcripts (DETs) among 60,660 transcripts involved in this study, many of which were novel. Further, the functions of this global transcript panel (including protein-coding transcripts, and lncRNAs) was assesed using a multiparametric approach.

Finally, lncRNA transcriptomic data was extracted from the RNA-seq dataset from the PC population, further characterizing 38 lncRNAs that were significantly differentially expressed, with most falling into the 125 DETs. To evaluate the lncRNA involvement, 4 ML algorithms were used to predict and distinguish between TMAs and TWAs. These algorithms included multivariate logistic regression (LR), support vector machine (SVM), random forest classifier (RFC), and eXtreme Gradient Boosting Classifier (XGBC). Several techniques were used to further reduce the bias within the included sample as described in the methodology.

Training and evaluation of the ML algorithms was performed by separating the dataset from the 38 DETs into a training set and a testing set to eventually evaluate the performance of each of the models. Out of all the ML algorithms, SVM and RFC were able to predict TMAs and TWAs with 76% accuracy using the 38 lncRNA data, suggesting important implications for the specified set of lncRNAs in PC. To the best of my knowledge, this is the first study to identify the involvement of this specific lncRNA panel in PC, with many novel lncRNAs lacking any studies performed on which.

The results of this research could potentially have important clinical implications, as the novelty of the identified lncRNAs requires further comprehensive validation and *in vitro* and *in vivo* investigations. The accuracy shown by the ML model suggests that these novel lncRNAs could be used as biomarkers and further targeted for improved diagnosis and outcome in PC patients.

## 2. Materials and methods

### 2.1. Data acquisition

TCGA database was used for data collection and is available at https://www.cancer.gov/tcga. Exploration of TCGA-PAAD project data to acquire pancreatic RNA-seq data was performed on 25/10/2023. File filters applied included a) Data Category: transcriptome profiling; b) Data Type: Gene Expression Quantification; c) Experimental strategy: RNA-Seq; d) Access: open. The case filters applied included the following: a) primary site: pancreas; b) project: TCGA-PAAD; and c) disease type: ductal and lobular neoplasms, adenomas and adenocarcinomas.

The inclusion criteria were that for each RNA-seq dataset to be of similar structure, for the predefined PC tumors mentioned in the filters, regardless of age and gender. Primary tumors, regardless of metastatic stage, were also included. Exclusion criteria included defects in dataset structure, RNA-seq for tumor adjacent tissues, or those that had undergone prior therapy to a potential previous malignancy. Records with annotations specifying that tumor data were incorrectly labeled in terms of whether the tumor was neoplastic, were also excluded.

Further categorization was performed for the acquired data using Excel sheets. For TNM subgroup analysis, tumors with staging data were categorized into tumors with metastatic activity, which included those classified as M1, MX/M0 and N1 or above, and tumors without metastatic activity, which included those classified as M0N0. Acquired data were also filtered to include only lncRNA gene expression quantification. This subgrouping was performed prior to DGEA to assess DETs between TWAs and TMAs.

### 2.2. Data analysis

Bioinformatics analysis was conducted on the data following matching the subjects to the study’s inclusion and exclusion criteria. Python v3.11 (available at https://www.python.org/) was used in an Anaconda jupyter lab environment ^14,15^. To restructure the dataset up for the study population RNA-seq datasets and to import the data into Python, the glob module was used ^16^. Data structure manipulation and organization was performed using pandas library v1.5.3 ^17^. Libraries such as numpy and scipy were also utilized for data processing ^18,19^.

Differential gene expression analysis (DGEA) was performed using PyDESeq2, an R package implemented in Python that has been suggested to be reliable and comparable to the R package^20^. The DETs were matched to gene symbols and further visualized using the matplotlib^21^, seaborn^22^, and sanbomics^23^ packages. PyDESeq2 calculates the significance of transcripts using the Wald test, performs count normalization using the trimmed mean of M values (TMM), similar to DESeq2, and relies on the statsmodels library ^24,25^. Using count normalization has been shown to have higher accuracy than TPM (transcripts per million) and FPKM (fragments per kilobase of transcript per million fragments mapped) ^26^. A more comprehensive description of the package is available elsewhere ^27^. Significant differentiation after adjustment of p values was considered at p<0.05 and an absolute log2-fold change (log2FC) of >0.5.

A heatmap of the DEGs was made through the matplotlib ^21^ package as well. Pearson’s correlation coefficient was calculated and mapped for all gene transcript data.

### 2.3. Gene set and ontology enrichment analysis

Gene set enrichment analysis (GSEA) is a method of interpreting gene-wide expression profiles^28^. GSEA was performed using the GSEApy v1.0.6 package, a Rust implementation of GSEA in python, used for performing computation of RNA-seq count data to evaluate predefined gene sets in association with different phenotypes. Gene expression data was ranked using the prerank function available in the package. The accuracy of this package has been previously proven, and the method to use it is described extensively elsewhere^29^.

Enrichment was performed for several gene collections from MSigDB available at (https://www.gsea-msigdb.org/) and miRTarBase 2017^30^. Gene sets and collections that were evaluated for enrichment were c2.cp.kegg.v2023.1.Hs.symbols, c3.mir.v2023.1.Hs.symbols, c3.tft.v2023.1.Hs.symbols, c4.cgn.v2023.1.Hs.symbols, c5.go.bp.v2023.1.Hs.symbols, c5.go.cc.v2023.1.Hs.symbols, c5.go.mf.v2023.1.Hs.symbols, c5.hpo.v2023.1.Hs.symbols, c6.all.v2023.1.Hs.symbols, h.all.v2023.1.Hs.symbols, and miRTarBase_2017.

Gene Ontology (GO) is a detailed resource with annotations of gene and gene product functions ^31,32^. It provides the potential to describe gene functions by assigning them to specific terms in which the transcripts’ genes are linked, detailing their relationships with each other. GO term enrichment was performed through GSEApy, and the results were extracted through tools available in said package.

The false discovery rate (FDR) was considered significant when FDR<0.05. Visualization of GSEA results was performed using tools from GSEApy. Data collected from GSEA results included terms, FDR, enrichment and negative enrichment scores, as well as matched genes. The minimum matching size for gene sets when performing GSEA for the global RNA-seq panel was set to 150. However, for the lncRNA panel, the minimum matching size was set to 3, as there were few enriched gene sets.

### 2.4. ML models

Multivariate LR, SVM, RFC, and XGBC were employed to predict metastatic risk for the population based on the lncRNA count data from TCGA. DETs were extracted from DGEA for use as sole predictors of metastatic progression in the study population. Analysis of the models’ accuracy was performed using packages from the scipy, scikit-learn, and matplotlib libraries.

To train the ML algorithms, data were categorized into a training set (70% of the data) and a testing set (30%). A random state number was set for all the implemented ML models to dictate a specific seed of randomness during the analysis to maintain reproducibility. For binary classification, TNM stage of IIa or below was designated “0” and considered the TWM for the ML algorithms, while TNM stage IIb or above was designated “1” and considered the TMA. The testing sets were hidden from the ML algorithms to evaluate the predictive capacity performance following model training.

Furthermore, hyperparameter tuning was performed to improve the predictive accuracy of the model. This was done through the GridSearchCV and BayesianSearchCV modules. Fivefold cross-validation was set as a parameter, and data regularization was done through L2 method, all of which have been shown to reduce bias and lower classification errors, also reducing sensitivity to outliers^33^. The inverse of the regularization strength (or penalty values) was set according to the optimal values found by the search modules specified above. To identify the best parameters, values were also tested over 50 iterations. Moreover, the synthetic minority oversampling technique (SMOTE) was performed to artificially increase TWM population numbers to reduce bias, which has proven to be a powerful tool in improving ML accuracy and addressing imbalanced samples ^34^.

These methods of standardization were performed for all ML algorithms used. ML algorithms used were also provided by the scikit-learn and XGBoost libraries. All of the algorithms consist of supervised machine learning algorithms, and are commonly used for classifications of tumors^35,36^. Further, L2 regularization has been considered to improve the accuracy of the ML algorithms ^37^.

To assess the performance of the ML algorithms, several evaluations were performed for each model. Accuracy is a very commonly used ML evaluation metric, here representing the ratio between the correctly determined TMAs and TWAs. Recall represents the sensitivity, describing the rate of correctly classified TMAs. Precision describes the ratio between correctly identified TMAs and all samples designated “TMA”. The F1 score metric represents the mean of precision and recall. A thorough description of the evaluation metrics used here is beyond the scope of this article, and has been explained comprehensively by Hicks et, al. elsewhere^38^. Area under the curve (AUC) was also used for evaluating the measures, and ML models having an AUC between 0.6 and 0.75 are considered to show possibly helpful discrimination (classification capacity), while above 0.75 indicate a clearly helpful classification capacity, as described elsewhere^39^.

## 3. Results

### 3.1. Primary characteristics of the study population

Of the 179 retrieved records, 23 were excluded for the following annotations: a) “This case is a neuroendocrine tumor and should not have been included in the PAAD study” (n = 8); b) “Per the PAAD EPC, this tumor is a normal pancreas with atrophy” (n = 5); c) “Per the PAAD EPC, this tumor is an atrophic pancreas” (n = 3); d) “Per the PAAD EPC, this tumor is a noninvasive IPMN” (n = 1); e) “Per the PAAD EPC, this tumor is an acinar cell carcinoma” (n = 1); f) “Per the PAAD EPC, this tumor is a normal ampula of Vater” (n = 1); g) “The PAAD EPC states that this case likely did not arise in the pancreas (ampullary)” (n = 1); h) “Systemic treatment given to the prior/other malignancy” (n = 1); i) “Per the PAAD EPC, this tumor is an atrophic pancreas with a single focus of low-grade PanIN” (n = 2); “Samples identified in the sample sheet with a sample type of “Solid Tissue Normal” (from normal tissue adjacent to malignancy)” (

According to the flow diagram found in **Figure 1**. A total of 151 patient records were included. **Table 1** summarizes the characteristics of the cohort. Notably, 115 records were classified as TMAs, while 36 were classified as TWAs. Of the TMAs, 116 were diagnosed as TNM stage IIb, and 8 were diagnosed as stage III and IV. For the TWAs, 26 were at TNM stage IIa.

**Figure 1.**
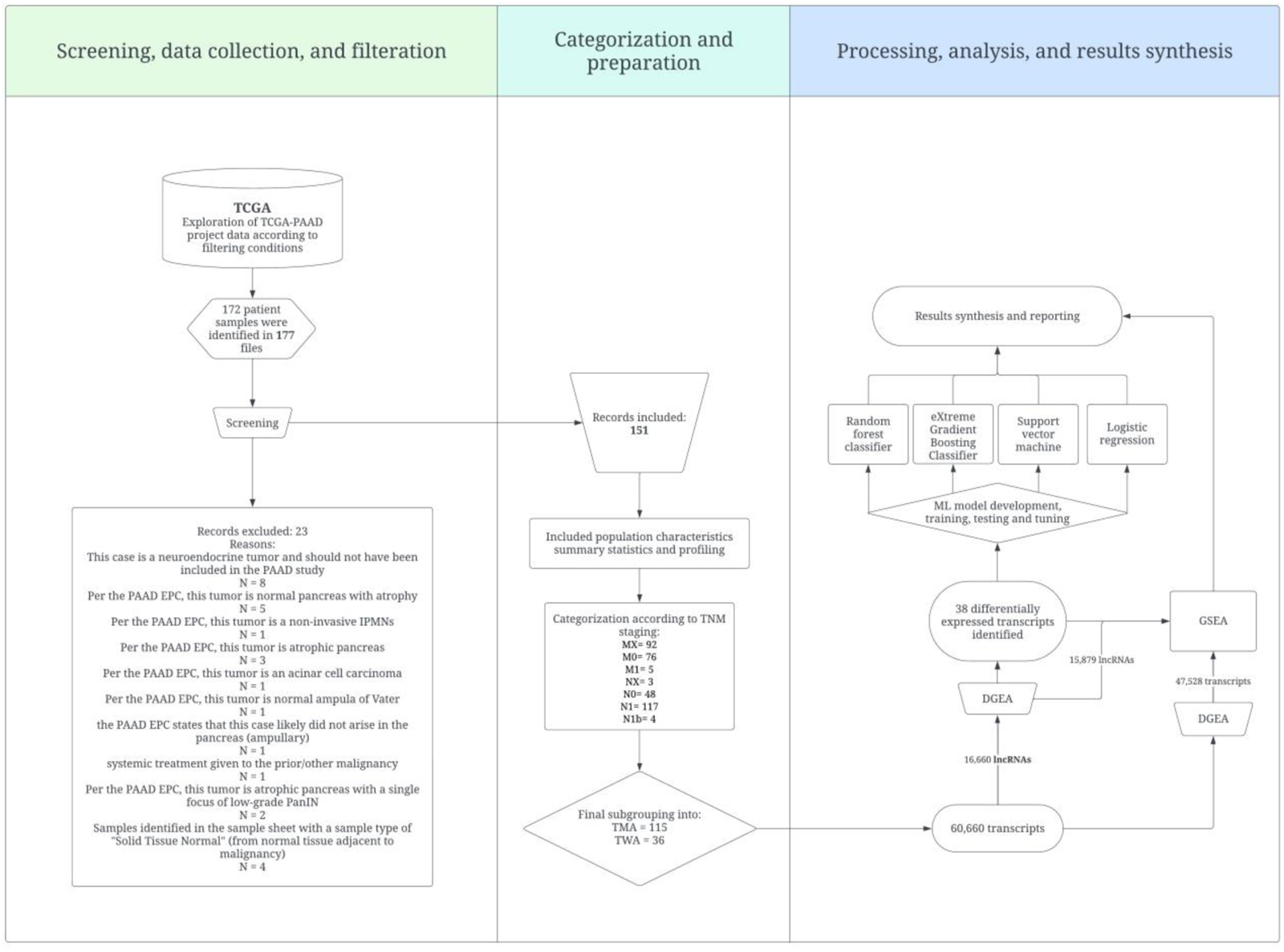
Flow diagram of the study. Created with Lucidchart, www.lucidchart.com. TCGA: The Cancer Genome Atlas; PAAD: Pancreatic adenocarcinoma; TMA: Tumor with metastatic activity; TWA: Tumor without metastatic activity; DGEA: Differential gene expression analysis; GSEA: Gene set enrichment analysis; ML: Machine learning.

**Table 1.** Population primary characteristics.

| Table 1. Population primary characteristics |  |
| --- | --- |
| General Characteristics |  |
| Average age | 64.62209 |
| Confidence | 1.173259 |
| STDEV | 10.92365 |
| Max age | 88 |
| Min age | 35 |
| Males | 80 |
| females | 70 |
| Pancreas, NOS | 14 |
| Head of pancreas | 112 |
| Body of pancreas | 11 |
| Tail of pancreas | 11 |
| Infiltrating duct carcinoma, NOS | 133 |
| Adenocarcinoma, NOS | 16 |
| **Included patient records characteristics** |  |
| TMA | 115 |
| TWA | 36 |
| MX | 78 |
| M0 | 68 |
| M1 | 5 |
| NX | 1 |
| N0 | 39 |
| N1 | 108 |
| N1b | 3 |
| **Staging** |  |
| I | 0 |
| III | 3 |
| IIb | 106 |
| IIa | 23 |
| IV | 5 |
| **STDEV**; Standard deviation; **NOS**: Not otherwise specified; **TMA**: Tumor with metastatic activity; **TWA**: Tumor without metastatic activity |  |

The age range of the total patient sample was between 35 and 88 years old (mean = 64.66 ± 10.91). Ninety-four were males, and 78 were females. When reported, 143 had infiltrating duct carcinoma, and 16 had adenocarcinoma as the primary diagnosis. Eight had neuroendocrine tumors but were excluded. Seventeen pancreatic tumors had no specified location, 125 were pancreatic head lesions, 15 were pancreatic body lesions, and 13 were pancreatic tail lesions.

The RNA-seq data included 60,660 transcript expression profiles for each of the included patient and control samples. Transcriptomic profiling was performed for the same genes in all patient samples. Of the available transcripts, 16,901 were lncRNAs. After removing lncRNAs with 0 values among all patients, 15,879 lncRNAs remained. All details regarding the included samples are available in **Supplementary Material 1.**

### 3.2. DGEA and GSEA of all transcripts

A total of 60,660 gene transcripts were filtered following PyDESeq2 analysis, and unavailable values were dropped, resulting in 47,528 transcripts. DGEA revealed 125 differentially expressed transcripts, as shown in **Table 2**, and the top DETs are shown in **Figure 2**. Notably, ADH7, SERPINB13, MIR205HG, NTS, and LINC01300 were the most downregulated DETs, with log2FC values of -3.42295, -3.4189, -3.12513, – 3.02808, and -2.72096, respectively. The most upregulated DETs were PAX7, AC010789.1, TMPRSS15, DEFA6, and DEFA5 and had log2FC values of 3.149596, 3.506053, 3.538356, 3.594891, and 4.800701, respectively.

**Figure 2.**
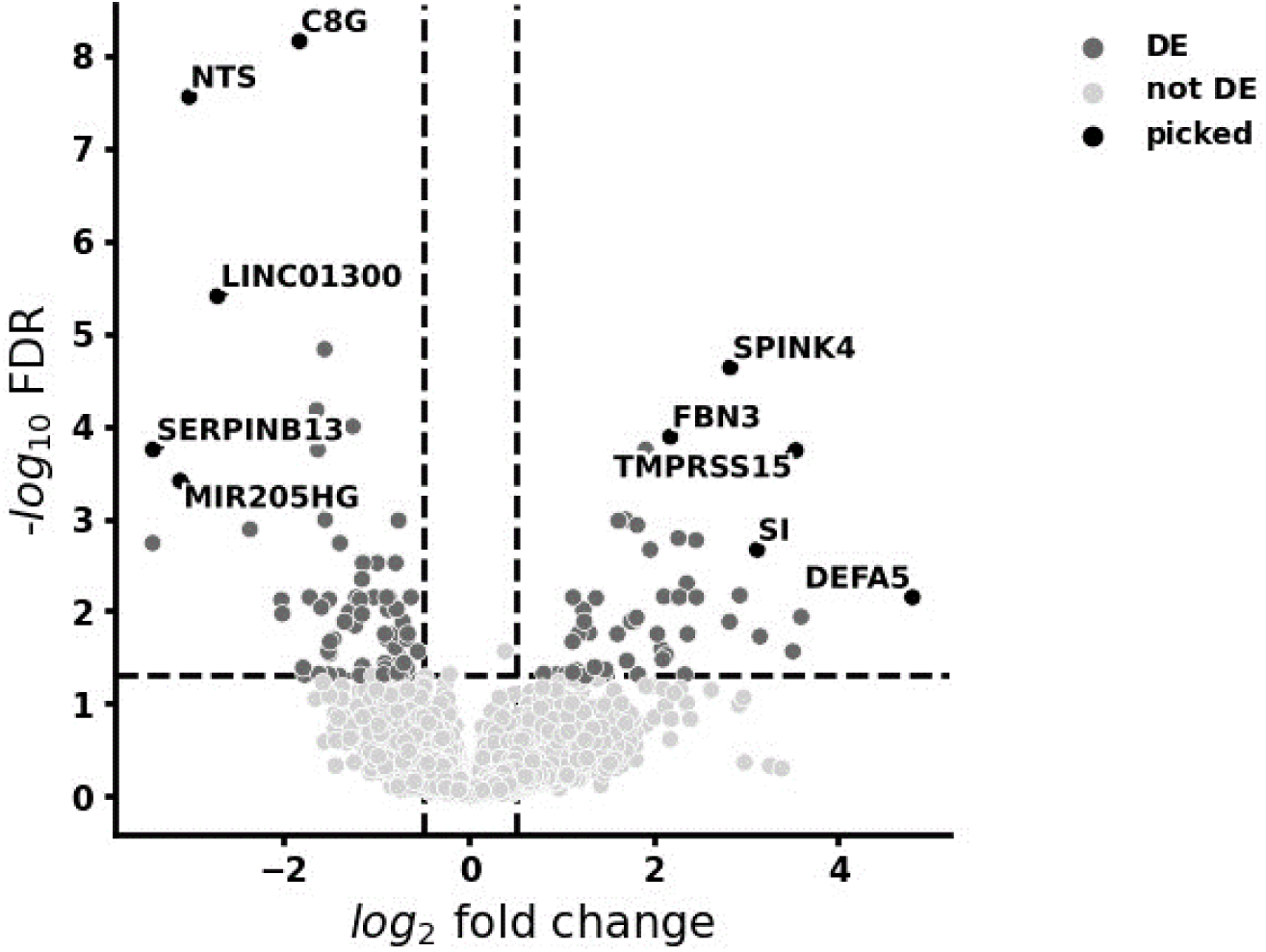
Differentially expressed transcripts in PC. Absolute log2FC>0.5 and adjusted p value<0.05 were considered as the significance thresholds.

**Table 2.** Differentially expressed protein-coding and long non-coding transcripts found in the global RNA-seq population.

|  |  |  |  |  |
| --- | --- | --- | --- | --- |
| ENSEMBL ID | Symbol | log2FoldChange | Rank | Adjusted p-value |
| ENSG00000196344 | ADH7 | -3.42295 | -4.80123 | 0.001852 |
| ENSG00000197641 | SERPINB13 | -3.4189 | -5.41464 | 0.00018 |
| ENSG00000230937 | MIR205HG | -3.12513 | -5.2119 | 0.000392 |
| ENSG00000133636 | NTS | -3.02808 | -7.0417 | 2.79E-08 |
| ENSG00000253595 | LINC01300 | -2.72096 | -6.20744 | 3.95E-06 |
| ENSG00000196427 | NBPF4 | -2.37012 | -4.89494 | 0.001312 |
| ENSG00000122133 | PAEP | -2.21459 | -5.13911 | 0.00054 |
| ENSG00000137975 | CLCA2 | -2.02911 | -4.3518 | 0.007622 |
| ENSG00000241794 | SPRR2A | -2.01833 | -4.2515 | 0.010747 |
| ENSG00000176919 | C8G | -1.83355 | -7.32595 | 6.96E-09 |
| ENSG00000285722 | AC207130.1 | -1.7953 | -3.80052 | 0.04115 |
| ENSG00000162951 | LRRTM1 | -1.77758 | -3.70984 | 0.049095 |
| ENSG00000075673 | ATP12A | -1.72492 | -4.41944 | 0.007122 |
| ENSG00000273143 | DUSP5-DT | -1.64896 | -5.64949 | 6.75E-05 |
| ENSG00000230916 | MTCO1P53 | -1.63588 | -5.3881 | 0.000181 |
| ENSG00000170477 | KRT4 | -1.62228 | -3.73908 | 0.047978 |
| ENSG00000258010 | AC016705.1 | -1.59722 | -4.30607 | 0.009204 |
| ENSG00000086570 | FAT2 | -1.58451 | -5.08612 | 0.00067 |
| ENSG00000214711 | CAPN14 | -1.56223 | -5.95983 | 1.48E-05 |
| ENSG00000101197 | BIRC7 | -1.55642 | -4.98114 | 0.001031 |
| ENSG00000110680 | CALCA | -1.52146 | -3.93287 | 0.02735 |
| ENSG00000130822 | PNCK | -1.51657 | -3.71624 | 0.04873 |
| ENSG00000166558 | SLC38A8 | -1.51515 | -4.3587 | 0.007531 |
| ENSG00000015592 | STMN4 | -1.50729 | -3.91404 | 0.02896 |
| ENSG00000205426 | KRT81 | -1.50078 | -4.00887 | 0.021539 |
| ENSG00000154975 | CA10 | -1.46245 | -4.02791 | 0.020145 |
| ENSG00000016602 | CLCA4 | -1.40473 | -3.69932 | 0.049966 |
| ENSG00000124466 | LYPD3 | -1.39559 | -4.79298 | 0.001855 |
| ENSG00000228705 | LINC00659 | -1.34601 | -4.17454 | 0.013082 |
| ENSG00000134339 | SAA2 | -1.29552 | -4.26585 | 0.010255 |
| ENSG00000108786 | HSD17B1 | -1.2613 | -6.32798 | 2.43E-06 |
| ENSG00000121552 | CSTA | -1.25807 | -5.5557 | 0.000101 |
| ENSG00000116014 | KISS1R | -1.23273 | -4.14973 | 0.014369 |
| ENSG00000204882 | GPR20 | -1.21769 | -4.39539 | 0.007122 |
| ENSG00000184564 | SLITRK6 | -1.17582 | -3.70529 | 0.049585 |
| ENSG00000253522 | MIR3142HG | -1.17478 | -4.3625 | 0.007531 |
| ENSG00000255129 | TTC12-DT | -1.15786 | -4.24636 | 0.01081 |
| ENSG00000233828 | MIR4280HG | -1.15767 | -4.56131 | 0.004522 |
| ENSG00000132746 | ALDH3B2 | -1.15254 | -3.81836 | 0.039435 |
| ENSG00000181652 | ATG9B | -1.14738 | -4.6546 | 0.003036 |
| ENSG00000115008 | IL1A | -1.12498 | -3.77886 | 0.043629 |
| ENSG00000177627 | C12orf54 | -1.02545 | -4.3938 | 0.007122 |
| ENSG00000180739 | S1PR5 | -0.99067 | -4.65304 | 0.003036 |
| ENSG00000272948 | AP001412.1 | -0.92415 | -3.72701 | 0.048613 |
| ENSG00000167971 | CASKIN1 | -0.90985 | -3.8412 | 0.036673 |
| ENSG00000278743 | AC087239.1 | -0.90938 | -4.0769 | 0.01772 |
| ENSG00000175189 | INHBC | -0.90458 | -3.79432 | 0.041787 |
| ENSG00000272906 | AL353708.3 | -0.88897 | -4.40552 | 0.007122 |
| ENSG00000178445 | GLDC | -0.88519 | -4.03342 | 0.020145 |
| ENSG00000268041 | ERFL | -0.8732 | -4.28133 | 0.009738 |
| ENSG00000254266 | PKIA-AS1 | -0.86706 | -4.38841 | 0.007122 |
| ENSG00000117407 | ARTN | -0.8143 | -4.08225 | 0.01772 |
| ENSG00000204963 | PCDHA7 | -0.79643 | -3.97143 | 0.024672 |
| ENSG00000286810 | AL513128.3 | -0.793 | -4.66358 | 0.003036 |
| ENSG00000268403 | AC132192.2 | -0.78124 | -4.29421 | 0.00953 |
| ENSG00000277218 | AL139123.1 | -0.77132 | -3.75731 | 0.046252 |
| ENSG00000102466 | FGF14 | -0.76382 | -3.83777 | 0.036813 |
| ENSG00000100162 | CENPM | -0.76299 | -3.89051 | 0.031239 |
| ENSG00000232573 | RPL3P4 | -0.76122 | -4.96717 | 0.00105 |
| ENSG00000237181 | PRKAR1B-AS1 | -0.75848 | -4.09162 | 0.017711 |
| ENSG00000233901 | LINC01503 | -0.73675 | -3.75264 | 0.046672 |
| ENSG00000267710 | EDDM13 | -0.71591 | -4.18507 | 0.013075 |
| ENSG00000196420 | S100A5 | -0.70536 | -3.75049 | 0.046672 |
| ENSG00000287575 | AL390755.3 | -0.70456 | -3.84387 | 0.036651 |
| ENSG00000227256 | MIS18A-AS1 | -0.66983 | -3.80296 | 0.041147 |
| ENSG00000263412 | NFE2L1-DT | -0.66855 | -4.03071 | 0.020145 |
| ENSG00000158292 | GPR153 | -0.6636 | -3.71762 | 0.04873 |
| ENSG00000270426 | AC099343.2 | -0.66056 | -4.09624 | 0.017609 |
| ENSG00000269961 | ERBIN-DT | -0.62896 | -4.4063 | 0.007122 |
| ENSG00000270659 | AC079610.1 | -0.55231 | -3.93098 | 0.02735 |
| ENSG00000109684 | CLNK | 0.811049 | 3.743661 | 0.047532 |
| ENSG00000007171 | NOS2 | 0.9501 | 3.727137 | 0.048613 |
| ENSG00000168004 | PLAAT5 | 1.046959 | 3.713844 | 0.04873 |
| ENSG00000217275 |  | 1.103167 | 3.945237 | 0.026898 |
| ENSG00000244675 | AC108676.1 | 1.121008 | 4.006455 | 0.021539 |
| ENSG00000249574 | AC226118.1 | 1.122255 | 3.758216 | 0.046252 |
| ENSG00000165186 | PTCHD1 | 1.127601 | 4.425499 | 0.007122 |
| ENSG00000204710 | SPDYC | 1.164409 | 3.774913 | 0.043911 |
| ENSG00000133317 | LGALS12 | 1.185546 | 4.074521 | 0.01772 |
| ENSG00000110195 | FOLR1 | 1.236702 | 4.28444 | 0.009738 |
| ENSG00000179766 | ATP8B5P | 1.248188 | 4.18952 | 0.013025 |
| ENSG00000243910 | TUBA4B | 1.26142 | 3.699883 | 0.049966 |
| ENSG00000231106 | LINC01436 | 1.269281 | 3.713818 | 0.04873 |
| ENSG00000079841 | RIMS1 | 1.301664 | 4.104695 | 0.017223 |
| ENSG00000254872 | LINC02688 | 1.359219 | 3.809804 | 0.040421 |
| ENSG00000077935 | SMC1B | 1.37522 | 4.3749 | 0.007278 |
| ENSG00000047936 | ROS1 | 1.462054 | 3.733667 | 0.048592 |
| ENSG00000250337 | PURPL | 1.478326 | 3.784362 | 0.043081 |
| ENSG00000211951 | IGHV2-26 | 1.607862 | 4.072645 | 0.01772 |
| ENSG00000113722 | CDX1 | 1.619806 | 4.955568 | 0.001058 |
| ENSG00000261409 |  | 1.673178 | 4.087027 | 0.01772 |
| ENSG00000095627 | TDRD1 | 1.695616 | 4.99184 | 0.001031 |
| ENSG00000275874 | PICSAR | 1.709471 | 3.859728 | 0.034709 |
| ENSG00000138823 | MTTP | 1.75384 | 4.194027 | 0.012975 |
| ENSG00000109182 | CWH43 | 1.779912 | 4.179552 | 0.013082 |
| ENSG00000286734 | AC133530.1 | 1.81407 | 4.219397 | 0.011788 |
| ENSG00000159251 | ACTC1 | 1.820261 | 4.924839 | 0.00118 |
| ENSG00000248635 |  | 1.82577 | 4.389008 | 0.007122 |
| ENSG00000124237 | C20orf85 | 1.830513 | 3.714347 | 0.04873 |
| ENSG00000070019 | GUCY2C | 1.911694 | 5.38153 | 0.000181 |
| ENSG00000185105 | MYADML2 | 1.961126 | 4.745671 | 0.002179 |
| ENSG00000179914 | ITLN1 | 2.039036 | 4.069503 | 0.01773 |
| ENSG00000130700 | GATA5 | 2.082396 | 3.959788 | 0.025605 |
| ENSG00000264404 | LINC02675 | 2.097143 | 3.871155 | 0.03347 |
| ENSG00000189052 | CGB5 | 2.11069 | 4.450157 | 0.006997 |
| ENSG00000198788 | MUC2 | 2.134906 | 3.90208 | 0.030102 |
| ENSG00000142449 | FBN3 | 2.180393 | 5.489955 | 0.000131 |
| ENSG00000250376 |  | 2.232802 | 4.650635 | 0.003036 |
| ENSG00000151365 | THRSP | 2.253533 | 4.419036 | 0.007122 |
| ENSG00000115850 | LCT | 2.267295 | 4.842754 | 0.001634 |
| ENSG00000198842 | STYXL2 | 2.278382 | 4.441746 | 0.007079 |
| ENSG00000205076 | LGALS7 | 2.337392 | 3.713775 | 0.04873 |
| ENSG00000166869 | CHP2 | 2.35771 | 4.533106 | 0.005018 |
| ENSG00000113196 | HAND1 | 2.366848 | 4.07674 | 0.01772 |
| ENSG00000091138 | SLC26A3 | 2.457377 | 4.823641 | 0.001724 |
| ENSG00000282122 | IGHV7-4-1 | 2.461323 | 4.384127 | 0.007122 |
| ENSG00000016490 | CLCA1 | 2.822016 | 4.177146 | 0.013082 |
| ENSG00000122711 | SPINK4 | 2.828123 | 5.854698 | 2.34E-05 |
| ENSG00000228674 | PPIAP59 | 2.932048 | 4.462678 | 0.006788 |
| ENSG00000090402 | SI | 3.119419 | 4.749043 | 0.002179 |
| ENSG00000009709 | PAX7 | 3.149596 | 4.052701 | 0.018812 |
| ENSG00000224817 | AC010789.1 | 3.506053 | 3.93045 | 0.02735 |
| ENSG00000154646 | TMPRSS15 | 3.538356 | 5.363727 | 0.000184 |
| ENSG00000164822 | DEFA6 | 3.594891 | 4.227086 | 0.011582 |
| ENSG00000164816 | DEFA5 | 4.800701 | 4.413106 | 0.007122 |

GSEA was subsequently performed, with libraries investigated available in **Supplementary Materials 2**. There were many gene sets enriched with the transcripts, as many transcripts were included in the study’s RNA-seq panel. Notably, several GO terms were enriched, as well as some terms from miRTarBase 2017, as shown in **Figure 3** **A and B**. FDR values were significant for the enriched terms (FDR<0.01).

**Figure 3.**
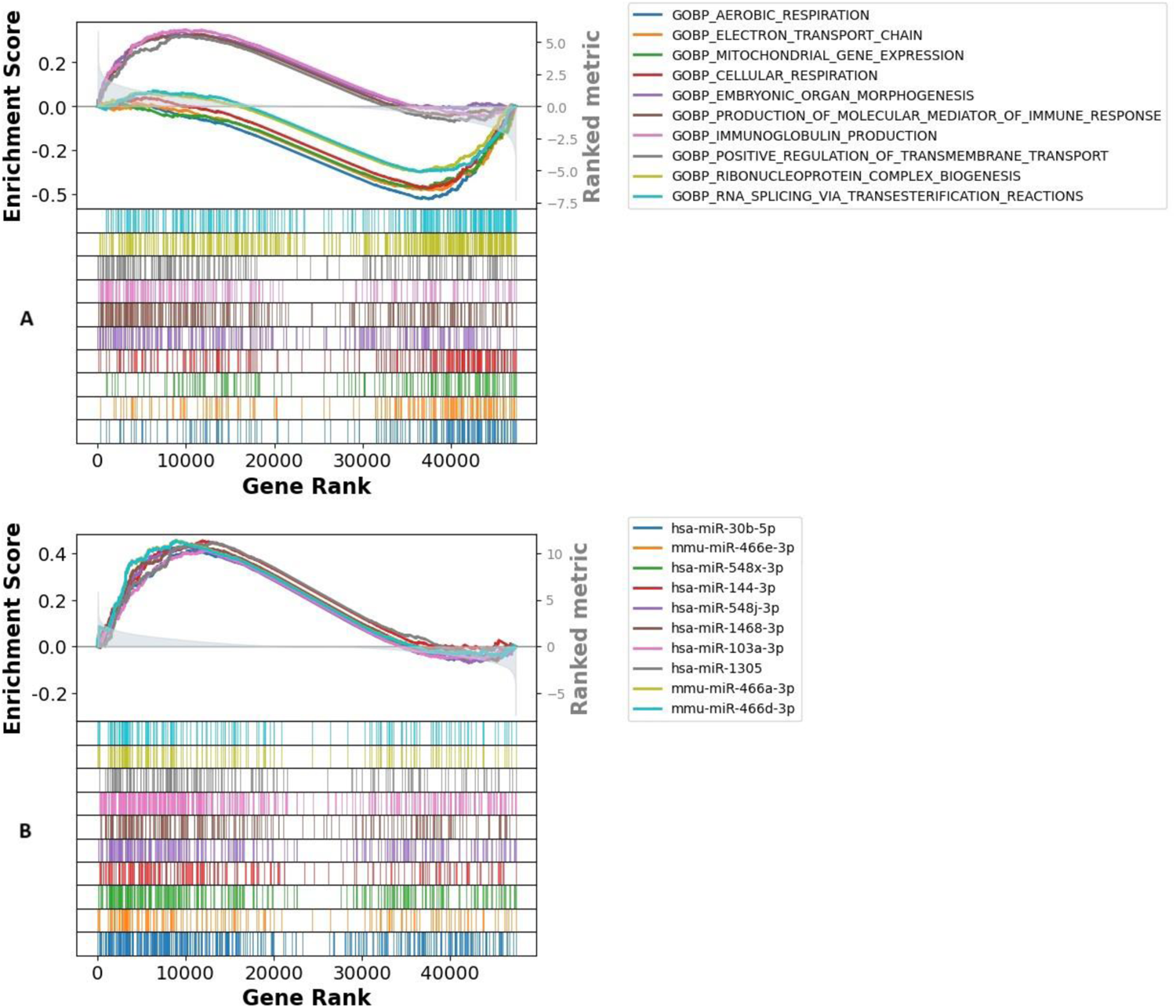
**A.** GOBP (GO biological process) term enrichment. Upregulated genes had a lower rank, and downregulated genes had a higher rank. The enrichment score correlates with the number of genes from the RNA-seq panel enriching the gene set with significantly differentiated expression. More transcripts enriching this term are downregulated in this study due to the enrichment score reaching – 0.5 since these genes have a higher density of higher ranked genes. **B.** miRTarBase_2017 term enrichment. Upregulated genes had a lower rank, and downregulated genes had a higher rank. The enrichment score correlates with the number of genes from the gene panel enriching the gene set with significantly differentiated expression. Here, the gene set was more enriched with the upregulated genes from the RNA-seq panel.

### 3.3. lncRNA DGEA, correlations, and GSEA

Further subgroup analysis was performed for lncRNAs in PC, which returned 16,901 expression values, for which PyDeseq2 was also used to analyze DETs. Dropping the 0-sum, duplicate, and unavailable values retrieved 15,568 lncRNAs. Of the lncRNA panel, 38 lncRNAs were significantly differentially expressed (shown in **Figure 4**).

**Figure 4.**
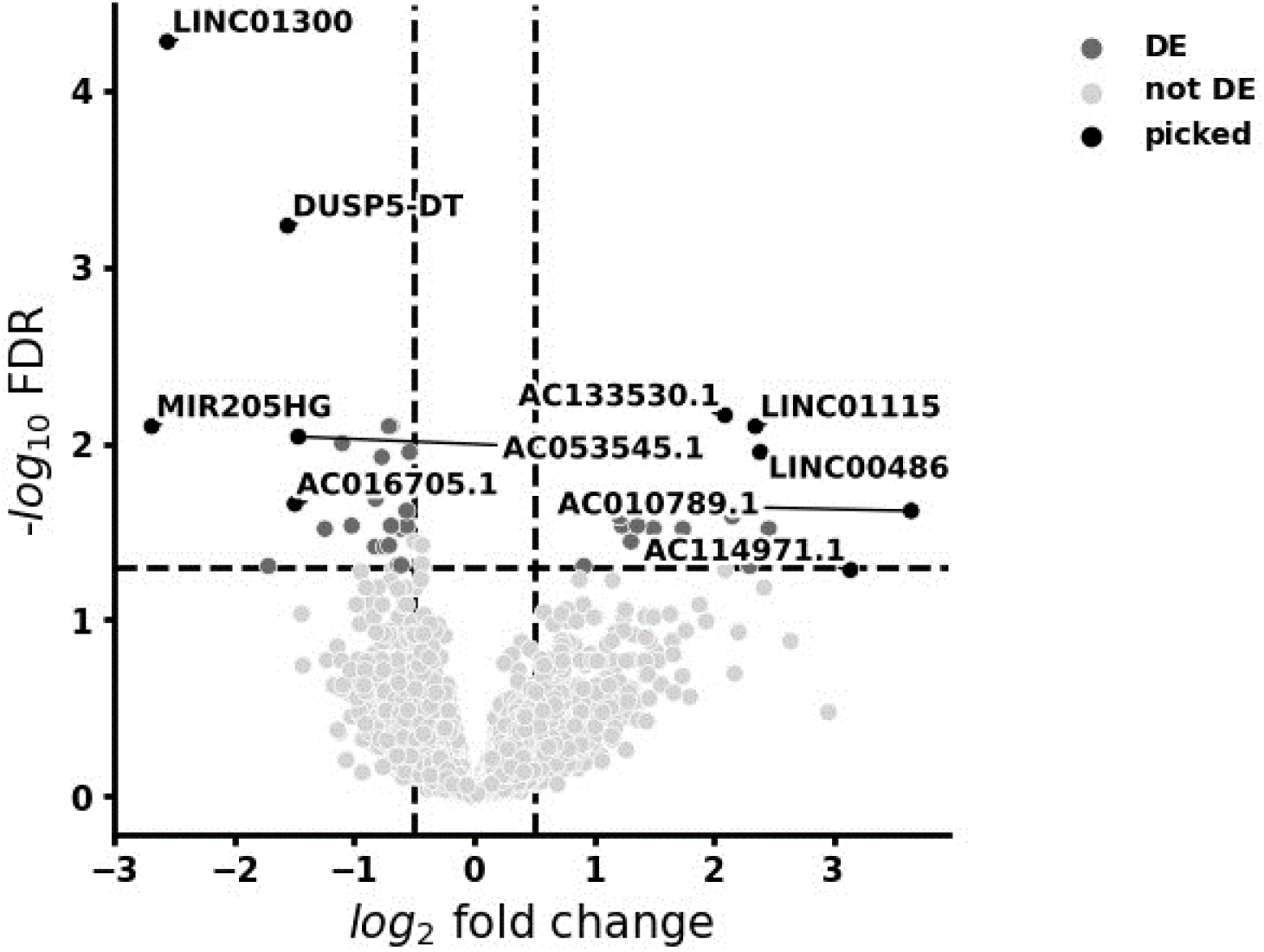
Differentially expressed lncRNA. Absolute log2FC>0.5 and adjusted p value<0.05 were considered as the significance thresholds.

Interestingly, the most downregulated lncRNAs were LINC01300, DUSP5-DT, AL513128.3, MIR205HG, and AC132192.2, with Log2FC values of -2.55682, -1.55378, -0.70877, -2.68894, and -0.68868, respectively. The most upregulated DET lncRNAs were AC010789.1, LINC00486, ENSG00000261409 (referred to as RF00019), LINC01115, and AC133530.1, with log2FC values of 2.154221, 1.214608, 3.647081, 1.705921, and 2.388161, respectively. Results of DGEA on the lncRNAs are shown in **Table 3**.

**Table 3.** DGEA of lncRNAs in PC.

| **Table 3. DGEA of lncRNAs in PC.** |  |  |  |  |
| --- | --- | --- | --- | --- |
| ENSEMBL ID | symbol | log2FoldChange | Rank | Adjusted p-value |
| ENSG00000253595 | LINC01300 | -2.55682 | -5.79229 | 5.25E-05 |
| ENSG00000273143 | DUSP5-DT | -1.55378 | -5.24801 | 0.000582 |
| ENSG00000286810 | AL513128.3 | -0.70877 | -4.56204 | 0.007988 |
| ENSG00000230937 | MIR205HG | -2.68894 | -4.5098 | 0.007988 |
| ENSG00000268403 | AC132192.2 | -0.68868 | -4.48214 | 0.007988 |
| ENSG00000287692 | AC053545.1 | -1.46588 | -4.42416 | 0.009158 |
| ENSG00000233828 | MIR4280HG | -1.09978 | -4.37975 | 0.00999 |
| ENSG00000269961 | ERBIN-DT | -0.53815 | -4.31064 | 0.011198 |
| ENSG00000272906 | AL353708.3 | -0.76963 | -4.27697 | 0.011946 |
| ENSG00000254266 | PKIA-AS1 | -0.82096 | -4.11833 | 0.020627 |
| ENSG00000258010 | AC016705.1 | -1.49399 | -4.08738 | 0.022009 |
| ENSG00000270426 | AC099343.2 | -0.56239 | -4.03685 | 0.024114 |
| ENSG00000253522 | MIR3142HG | -1.02076 | -3.95022 | 0.029262 |
| ENSG00000263412 | NFE2L1-DT | -0.5603 | -3.93248 | 0.029262 |
| ENSG00000277218 | AL139123.1 | -0.69149 | -3.90873 | 0.029262 |
| ENSG00000227256 | MIS18A-AS1 | -0.61208 | -3.88033 | 0.03036 |
| ENSG00000228705 | LINC00659 | -1.24398 | -3.8544 | 0.030461 |
| ENSG00000285886 | AC211476.6 | -0.70691 | -3.76746 | 0.037816 |
| ENSG00000272948 | AP001412.1 | -0.82452 | -3.75478 | 0.038514 |
| ENSG00000278743 | AC087239.1 | -0.74556 | -3.74815 | 0.038514 |
| ENSG00000285763 | AL358777.1 | -0.60768 | -3.67036 | 0.048983 |
| ENSG00000276791 | AC092117.1 | -0.63072 | -3.66641 | 0.048983 |
| ENSG00000285722 | AC207130.1 | -1.71493 | -3.65656 | 0.049573 |
| ENSG00000265933 | LINC00668 | 2.298864 | 3.643841 | 0.049573 |
| ENSG00000268388 | FENDRR | 0.916365 | 3.644596 | 0.049573 |
| ENSG00000231106 | LINC01436 | 1.30986 | 3.795489 | 0.035968 |
| ENSG00000248740 | LINC02428 | 2.458109 | 3.852876 | 0.030461 |
| ENSG00000275874 | PICSAR | 1.742661 | 3.862257 | 0.030461 |
| ENSG00000250337 | PURPL | 1.495827 | 3.887343 | 0.03036 |
| ENSG00000228709 | LINC02575 | 1.235504 | 3.908659 | 0.029262 |
| ENSG00000254872 | LINC02688 | 1.363374 | 3.920829 | 0.029262 |
| ENSG00000264404 | LINC02675 | 2.154221 | 3.993045 | 0.025979 |
| ENSG00000249574 | AC226118.1 | 1.214608 | 3.998806 | 0.025979 |
| ENSG00000224817 | AC010789.1 | 3.647081 | 4.04573 | 0.024114 |
| ENSG00000261409 |  | 1.705921 | 4.224935 | 0.013912 |
| ENSG00000230876 | LINC00486 | 2.388161 | 4.312419 | 0.011198 |
| ENSG00000237667 | LINC01115 | 2.347361 | 4.509832 | 0.007988 |
| ENSG00000286734 | AC133530.1 | 2.092716 | 4.689599 | 0.006905 |

Moreover, since the number of DETs was feasible, to further visualize the relationship between these lncRNAs, each transcript’s natural logarithm of 1 plus (normalized count) data was correlated to their respective PC cases, and Pearson’s correlation coefficients for all the lncRNAs were extracted. The results are visualized in **Figure 5**. A table of all Pearson’s correlation coefficients can be found in **Supplementary Material 3**.

**Figure 5.**
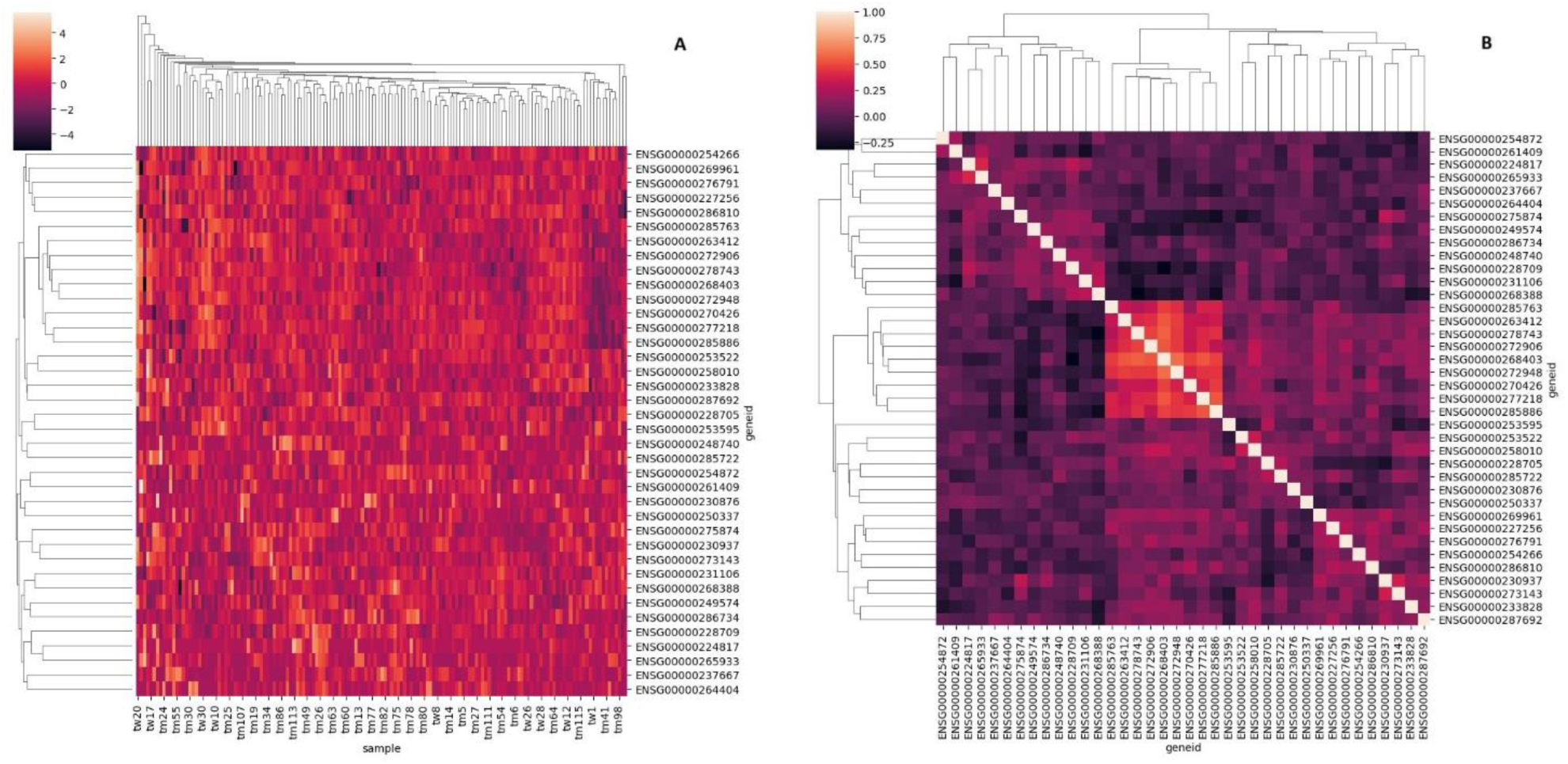
**A.** Hierarchical clustering heatmap of identified lncRNAs and their corresponding expression per PC case. The color gradient in the legend refers to the natural logarithm of 1 plus (normalized count) values. **B.** Hierarchical clustering heatmap of lncRNAs amongst the sample population. The color gradient in the legend refers to Pearson’s correlation coefficient. The dendrogram linkage is based on the correlation strength. Geneid: ENSEMBL ID for the gene encoding the respective lncRNA transcript. tw: TWAs; tm: TMAs.

GSEA and GO analyses were subsequently performed for all the lncRNA data. Due to the lack of studies on the genes of these transcripts, there was no significant enrichment in most databases. Notably, a few terms were enriched from the MSigDB c3.tft.v2023.1.Hs.symbols collection, which is focused on transcription factors. The results of the term enrichment for the top 10 terms in this collection are shown in **Figure 6**, and the results for insignificant term enrichment for other collections and databases can be found in **Supplementary Material 3.**

**Figure 6.**
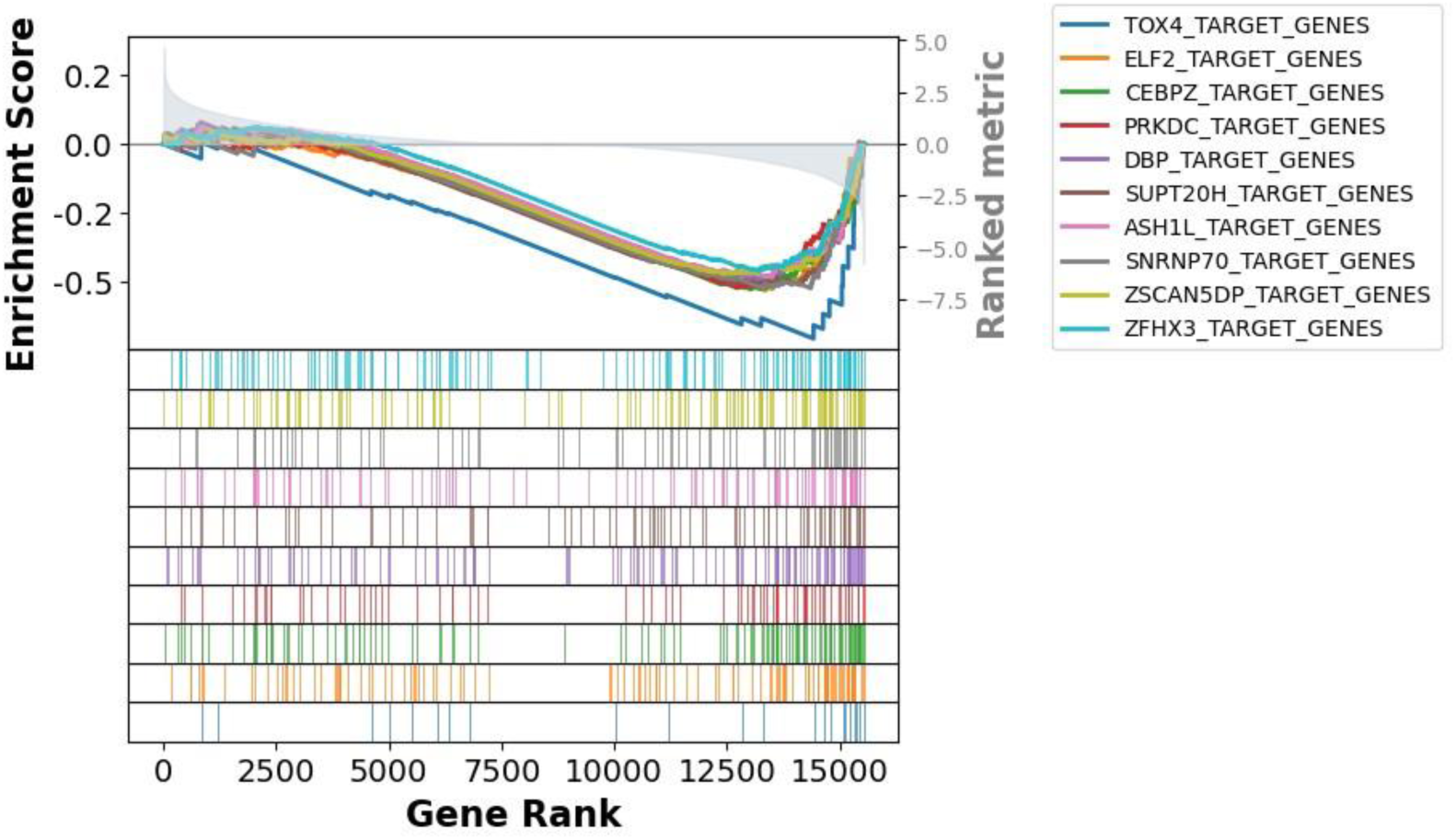
GSEA of lncRNAs in MSigDB transcription factors gene set. Terms are more significantly enriched with downregulated genes.

### 3.4. ML model prediction of PC metastatic potential according to lncRNA expression

Following the training and testing of each of the ML models, optimizations were performed to find the highest possible accuracy obtainable while reducing bias. Therefore, SMOTE was implemented in all the ML algorithms. Reducing sample imbalances improved the predictive accuracy of the utilized algorithms.

Following SMOTE implementation and thorough hyperparameter tuning, LR demonstrated an accuracy score of 73.91% when distinguishing between TMAs and TWAs when tested, as well as an F1 score of 82.57% and a recall of 90.63%. Regardless, the AUC for LR was 0.63. **Figure 7** **A and B** show the receiver operating characteristic (ROC) curve and for logistic regression following the implementation of SMOTE and the precision-recall (PR) curve.

As the LR model was the only allowing the determination of prediction coefficients, assessment of which lncRNA had the highest weight in predictions was performed. The most notable lncRNAs with positive correlation coefficients (>0.50) include: LINC02575, LINC01115, LINC02428, PURPL, and AL035425.3. While the most notable ones with negative correlations (<-0.50) indicating a lower likelihood for metastatic PC include: AC207130.1, and AL358777.3. **Figure 7** **C** shows the weight of each lncRNA (feature) in assisting the regression model in classifying test cases into TMAs and TWAs.

**Figure 7.**
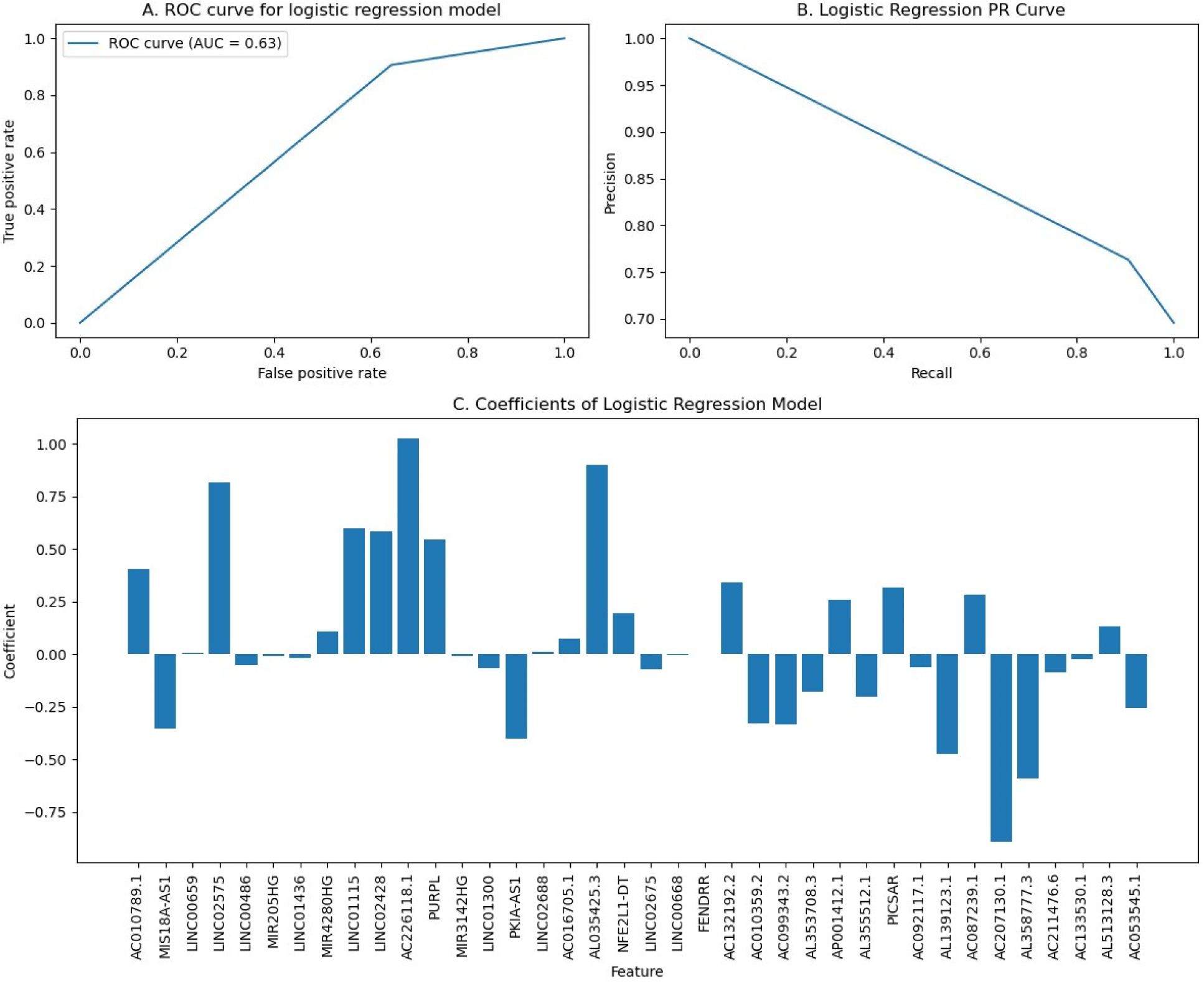
**A.** The LR model showed an AUC = 0.63, demonstrating relatively helpful classification performance, with good accuracy of detecting PC cases at TNM stage IIb or above. **B.** LR model accuracy of predicting positive values in comparison to the true positive rate (recall). **C.** Weights of each of the differentially expressed lncRNAs allowing the LR model to differentiate between non-metastatic tumors and metastatic tumors.

For the SVM model, SMOTE implementation, and hyperameter tuning also improved the predictive potential of the algorithm, which, on testing, returned an accuracy of 76.09%, with a true positive rate of 84.51% and a recall of 93.75%. **Figure A and B** show the ROC curve as well as the PR curve of the SVM model.

**Figure 8.**
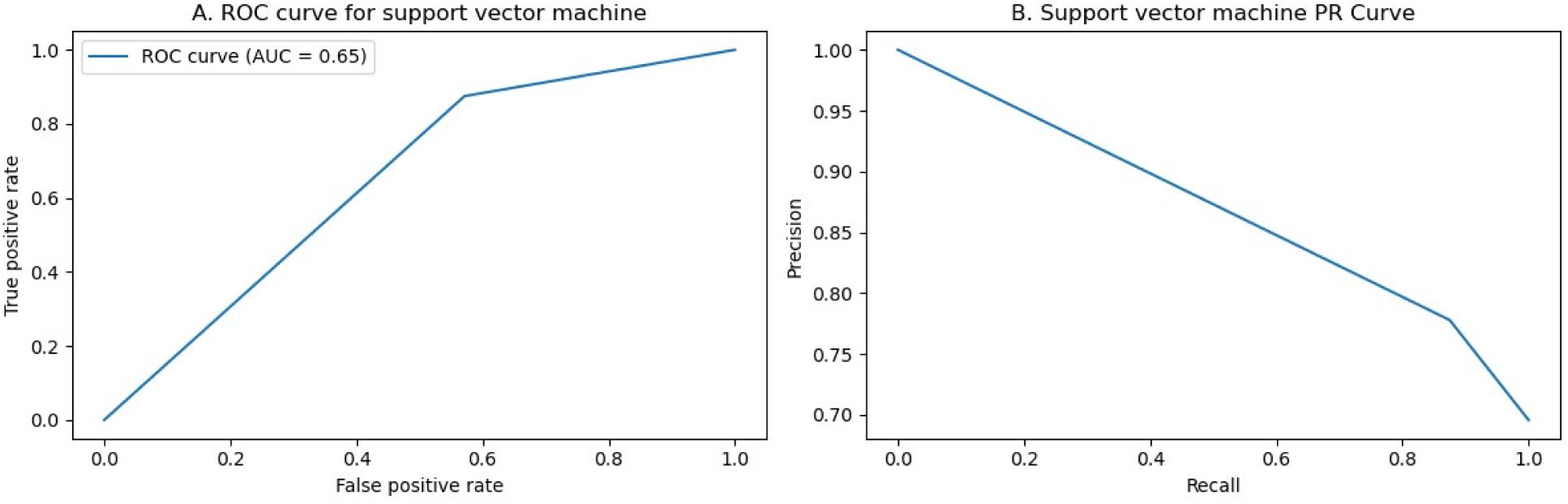
**A.** The SVM algorithm showed an AUC = 0.65, demonstrating modest performance when accurately detecting PC cases at TNM stage IIb or above and distinguishing them from less metastatic stages. **B.** SVM model accuracy of predicting positive values in comparison to its recall capacity.

RFC was one of the most accurate models; after hyperparameter tuning, it returned an accuracy of 76.09% and an F1 score of 81.96%, with a recall of 78.13%. Most importantly, the AUC for this model was 0.75, showing good performance in classifying the tumors. Regardless, the lncRNA panel consisting of 38 differentially expressed lncRNAs allowed the ML algorithms to discern advanced TNM stages from relatively early TNM stages in PC. **Figure 9** **A and B** also show the RFC model accuracy and PR curve.

**Figure 9.**
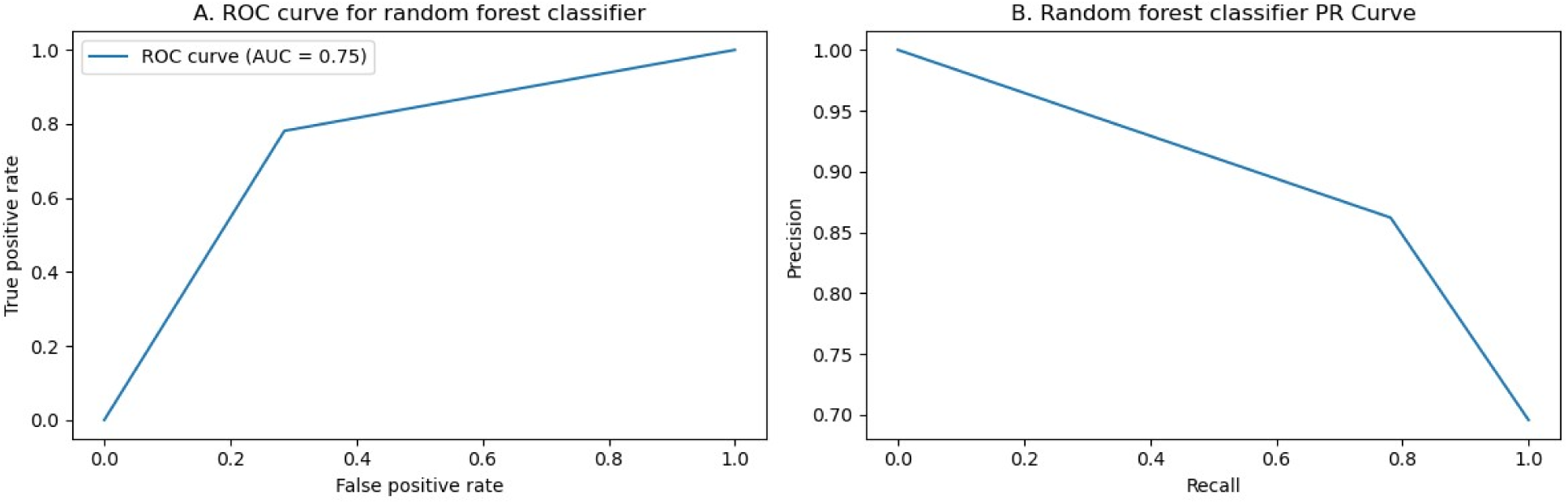
**A.** The RFC ROC AUC was 0.75, demonstrating clearly significant classification accuracy of detecting PC cases among the other ML algorithms when using the differentially expressed lncRNA counts data. **B.** The RFC PR curve showed good recall, and acceptable precision.

As for XGBC, the model showed 71.73% accuracy; This specific model had the most inconsistency in predicting tumor types following each randomization. **Figure 10** **A and B** show the low AUC and its PR curve. Data regarding the evaluation of the ML algorithms are available in **Supplementary Material 4**.

**Figure 10.**
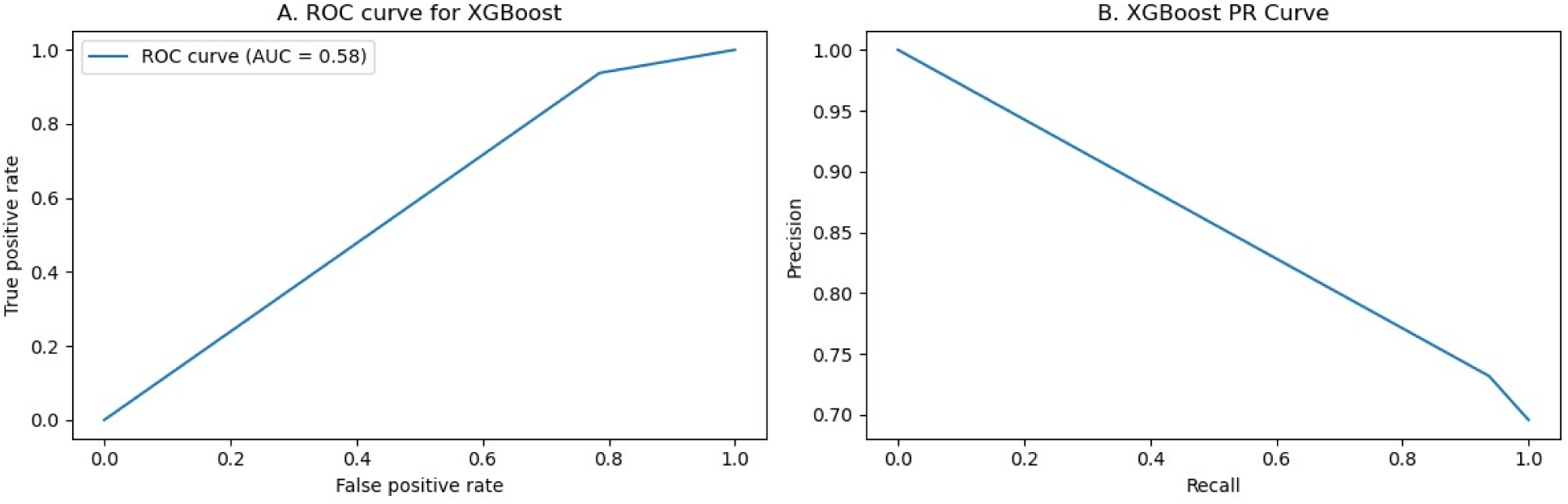
**A.** XGBC showed the lowest AUC of 0.58. While the accuracy for detecting metastatic PC cases was high, the false positive rate was also high. **B.** Poorest reliability amongst the ML algorithms in the XGBC model.

## 4. Discussion

Despite advances in diagnostics and therapeutics, PC remains a very challenging condition to treat, with consistently high mortality rates and limited available treatments^40,41^. Recently, research has focused on identifying prognostic markers for PC, and preclinical studies have identified several prognostic lncRNA signatures^8,42–44^. LncRNAs have been further suggested to have implications in diagnosis, drug resistance, and therapeutics in PC^4^. However, as most patients are often diagnosed at advanced stages of disease, mutational burdens show complex relationships with lncRNA regulation^4^. Therefore, as the literature suggests, these relationships must be investigated to adjust treatment modalities. This becomes even more crucial in the latter stages of PC.

This study aimed to provide details regarding DETs in PC first and then to further analyze differentially expressed lncRNA and assess the diagnostic potential of these lncRNAs during the transition from stage IIa and stage IIb and above. These lncRNAs were extracted after performing DGEA to extract 38 gene transcripts from the global RNA-seq panel among 151 patient samples. The diagnostic potential of lncRNAs was assessed using supervised ML techniques to predict metastatic transition. Four ML techniques with established accuracy in prediction were used in this research: LR^45^, SVM^46^, RFC^46^ and XBGC^47^.

DGEA of the global RNA-seq panel revealed 125 DETs, many of which were previously uninvestigated. Of the downregulated DETs, ADH7 was hypothesized to have implications when mutated in pancreatic injury^48^. NTS was also associated with PC^49^. However, SERPINB13 and MIR205HG were previously unexplored in PC but had been discussed in other cancers and were implicated in poor clinical outcomes^50,51^. No studies are available regarding LINC01300, which warrants further investigation. For the upregulated DETs, PAX7 was previously reported to have some relationship with cancers, yet studies regarding this specific gene transcript are lacking ^52^. For DEFA6 and DEFA5, a report suggested a link between them and clinical outcomes in colorectal cancer^53^. While there were no studies regarding AC010789.1 and TMPRSS15 in PC, some studies linked the potential implications of these transcripts with other cancers^54,55^.

GSEA for the global RNA-seq panel revealed several enriched pathways in many gene sets. For example, GO enrichment revealed that the RNA-seq panel significantly enriched pathways relevant in the regulation of aerobic respiration (GO:1903715), electron transport carrier chain (GO:0022900), and mitochondrial gene expression and translation into RNA transcripts (GO:0140053). Notably, of the miRTarBase enriched pathways, mir-30b-5p microRNA (miRNA) was previously linked to PC^56,57^. While miR-548x-3p has not been studied regarding its function in cancer, miR-144-3p was previously implicated in PC^58,59^. Additionally, mir-548j-3p had no studies documenting its relationship with cancer. For miR-1468-3p, some studies have suggested it as a biomarker for non-small cell lung cancer and prostate cancer^60,61^.

Following the filtering of the global RNA-seq panel to lncRNAs exclusively, DGEA revealed 38 differentially expressed lncRNAs, many of which were novel. LINC01300 and MIR205HG, as previously described, in addition to DUSP5-DT and AL513128.3, had no studies in PC, with the latter two completely lacking any studies on which. In contrast, one report regarding AC132192.2 indicated its relevance in prostate cancer^62^. For the upregulated lncRNAs, AC010789.1, as previously stated, had a report regarding its function in colorectal cancer^55,63^. LINC00486, RF00019, LINC01115, and AC133530.1 all lack validation studies in PC, but other reports indicate involvement in several diseases, including cancer^64–67^.

As these novel lncRNAs lack studies regarding their functions, GSEA of the selected MSigDB collections returned no significant enrichment but in one transcription factors collection. Notably, the most enriched pathway described genes containing one or more binding regions for a transcription factor that regulates cell fate and controls cell cycle progression from the mitotic phase to interphase, known as TOX high mobility group box family member 4 (TOX4)^68,69^. Interestingly, lncRNAs enriching this term were primarily downregulated.

To further explore the significance of the identified 38 lncRNAs, ML algorithms were employed to predict the metastatic state of cancer (designated “0” for stages IIa or below and “1” for stages IIb and above). The LR model suggested that a few lncRNAs may have more significance in metastatic progression, most notably: LINC02575, LINC01115, LINC02428, PURPL, AL035425.3, AC207130.1, and AL358777.3. All of which lacking studies in PC. Nonetheless, LINC02575 has been found to be implicated in proliferation of laryngeal squamous cell carcinomas ^70^; PURPL was indicated to have an involvement in ovarian and gastric cancers ^71–73^; and AL035425.3 was suggested to be implicated in the prognosis of triple negative breast cancer^74^.

Of all the ML algorithms, RFC showed superior accuracy to the other algorithms, showing an AUC of 0.75 and an accuracy of over 76%. RFC models have been previously shown to have superior performance to several other ML algorithms^39^. While there is much to be understood regarding the functions of the identified lncRNA panel, the accuracy shown by RFC reveals important aspects about the involvement of these lncRNAs in PC. These finding warrants further *in vitro* and *in vivo* investigations of the identified lncRNA panel.

For most of the identified lncRNA panels, this was the first study to uncover their involvement in PC. Regardless, there are many clinical implications for the findings discussed here. The results of this study suggest that the identified lncRNAs could be further utilized to assess the metastatic potential of PC, as well as aid in drug development, since these lncRNAs can be used as drug targets. Since their involvement allowed the prediction and distinction between TNM stages, further investigation of their functions seems crucial.

Despite the significant findings, this study is not without limitations. First, DEGA was performed for a large number of data, which likely raised data noise. Second, TWAs used as controls were low in number, as most samples had a stage IIb diagnosis, and SMOTE was necessary to utilize for the ML algorithms to reduce bias. Third, there was a lack of normal tissue control samples, which makes it difficult to provide more accurate assessments of the nature of these lncRNAs. Last, there might have been biases in the TCGA data from incorrect measurements or sequencing, potentially skewing the results of the RNA-seq data. All of these findings indicate that the findings of this study should be further validated and interpreted with caution.

Regardless, the presence of some evidence regarding some of the identified novel lncRNAs in other cancers suggests their potential involvement in PC proliferation and metastasis. This further adds to the implications of the findings discussed here and the importance of future research to address these novel lncRNAs as potential markers of metastatic progression in PC.

## 5. Conclusion

DGEA utilized in this study identified a set of 38 novel lncRNAs that could contribute to metastatic progression in PC. GSEA was unable to provide sufficient information to further describe the functions of these lncRNA, due to the scarcity of available data relevant to the transcripts identified. Since different ML algorithms were able to predict metastatic PC with acceptable accuracy and the RFC model predicted PC with 76% accuracy based on the 38 lncRNA panel, it is likely that these DETs participate in the metastatic progression of PC, warranting further investigation.

The significance and importance of this study is represented by the identified novel lncRNA set. Metastatic PC lacks sufficient studies regarding the involvement of lncRNAs in tumor proliferation and progression, especially those that use ML algorithms with proven accuracy. This is the first study of its kind to use this methodology to reveal the discussed lncRNA panel in PC to distinguish between early-stage and advanced PC. Regardless, more studies are needed to identify the role these genes play in PC metastasis and other cancers.

Based on the findings of this study, I suggest further research to take place into the roles of these RNAs in metastatic PC. *In vitro* and *in vivo* experiments must be conducted to further elucidate the functions these lncRNAs may take part in. The accuracy of the ML algorithms when classifying metastatic PC reveals that these lncRNAs could have important potentials in improving the diagnostic accuracy for metastatic PC when implemented with other techniques, and should be evaluated for therapeutic potentials.

## Data Availability

All raw data acquired from TCGA, in addition to all analyses performed on said data and source code utilized to perform the analyses mentioned in the methodology are available at the link https://github.com/hasanalsharoh/PanC.

## Notes

### Competing Interest Statement

The authors have declared no competing interest.

### Funding Statement

This study did not receive any funding.

### Author Declarations

All data used in this study was acquired from the cancer genome atlas (TCGA) available from https://portal.gdc.cancer.gov/projects/TCGA-PAAD. The search algorithm for retrieving the data can be provided on request. The filters picked included only data with open access. Data with controlled access was excluded.

### Summary of Updates

Figures 2 and 4 show a significance threshold of 0.75, instead of 0.5 as written in the description. Figure 5 description is for figure 5B, this issue happened as I was unsure of which of the figures to include due to size constraints. Nonetheless, in this revision, I include both figures (A and B) for clarity. Further revisions were done for calculations and everything else, results remain unchanged and conclusions unaltered thereof. Manuscript text remains unaltered following my revisions.

